# Echo intensity is not an accurate surrogate marker of intramuscular fat and fibrosis: Systematic review with meta-analyses

**DOI:** 10.64898/2026.09.19.26363379

**Authors:** Matheus D. Pinto, João Pedro Nunes, Ronei S. Pinto

## Abstract

**Background:** Intramuscular fat and fibrous tissue deposition increase with ageing, disuse, and disease and contribute to impaired muscle function and reduced ‘muscle quality’. Magnetic resonance imaging (MRI) and biopsy provide reference measures of muscle composition, but their cost, invasiveness, and limited accessibility restrict routine use. B-mode ultrasonography has emerged as a practical, low-cost alternative, with ultrasound-derived echo intensity widely interpreted as a surrogate measure (index) of intramuscular fat, fibrosis, and ‘muscle quality’. However, such interpretation warrants further scrutiny.

**Aim:** To quantify the relationship between ultrasound-derived echo intensity and reference measures of intramuscular fat and fibrous tissue and determine whether these relationships vary across muscles or according to anatomical and methodological factors.

**Methods:** A systematic search of six databases was conducted (last search: August 2026). Human and animal studies reporting correlations between echo intensity and measures of intramuscular fat or fibrous tissue using MRI, histology, or chemical analysis were synthesised using multilevel random-effects meta-analysis with study-level cluster-robust inference. Correlations were Fisher *z*-transformed and back-transformed for interpretation. Exploratory analyses examined fat compartment, muscle, muscle architecture, health status, study model, and distance-correction procedures.

**Results:** Twenty-eight studies met the eligibility criteria. For intramuscular fat, 204 effects from 23 independent studies yielded a weak-to-moderate pooled correlation with echo intensity (r=0.54, 95% CI: 0.45-0.61; 95% PI: −0.02-0.84; P<0.01), with substantial heterogeneity (I²=74.5%). The wide prediction interval suggests the true correlation could range from negligible to strong. No significant moderation effects were observed for study model (animal vs. human: r=0.46 vs. 0.55; P=0.26), health status (clinical vs. healthy: r=0.64 vs. 0.49; P=0.11), probe-to-muscle distance correction (corrected vs. raw echo intensity: r=0.57 vs. 0.53; P=0.25), or muscle architecture (non-pennate vs. pennate: r=0.63 vs. 0.53; P=0.56), although some categories contained few studies and effects. Model-based inference indicated a significant moderation effect for lipid compartment (intramyocellular lipid vs. total intramuscular fat vs. extramyocellular lipid: r=0.10 vs. 0.57 vs. 0.49; P<0.001) but this was not significant under cluster-robust inference (P=0.50). For fibrous tissue, six effects from four studies yielded a weak, non-significant pooled correlation (r=0.34, 95% CI [−0.39, 0.81], 95% PI [−0.85, 0.96]; P=0.24).

**Conclusions:** Echo intensity is weakly-to-moderately associated with intramuscular fat, and this varied substantially across studies and muscles, whereas evidence for intramuscular fibrous tissue is limited and uncertain. These findings provide limited support for interpreting echo intensity as a surrogate measure or an index of intramuscular fat or fibrosis or, on this basis, as an index of ‘muscle quality’.

## 1 BACKGROUND

Loss of muscle strength in ageing, disuse, and neuromuscular disorders reflects not only skeletal muscle atrophy but also changes in muscle composition that result in increased intramuscular fat infiltration and fibrous tissue deposition [1–3]. These ‘non-contractile’ tissue changes impair skeletal muscle contractile function [3], contribute to functional decline [4], and are associated with metabolic dysfunction and disease progression [2, 5]. Accurate quantification of intramuscular fat and fibrous tissue is therefore important for characterising muscle morphology and function and for monitoring disease progression [6].

Several techniques exist for assessing intramuscular fat and fibrous tissue in skeletal muscle [7]. Muscle biopsy provides detailed histochemical and structural information and is the reference method for evaluating fat and fibrous tissue content [8, 9], but its invasiveness, cost, and limited sampling area restrict its use in clinical and research settings. Magnetic resonance imaging (MRI) is the gold standard non-invasive method [6, 10, 11] and enables precise [6] and validated [12] separation and quantification of fat and water signals within muscle. However, MRI is also limited by high costs and restricted accessibility. These limitations have driven significant interest in other imaging techniques, with B-mode ultrasonography emerging as a practical, cost-effective, and portable alternative [13–15].

Ultrasound-derived muscle echo intensity is often considered a surrogate marker of ‘muscle quality’ that is thought to reflect intramuscular fat infiltration and connective/fibrous tissue [15]. Echo intensity is derived from the mean greyscale pixel intensity within a defined muscle region of interest within the B-mode image [16]. Higher values of echo intensity (brighter images) are thought to indicate greater intramuscular fat and fibrous tissue and reflect poorer ‘muscle quality’, whereas lower values (darker images) are assumed to represent higher proportions of contractile muscle tissue and water and to indicate better ‘muscle quality’. These interpretations stem from early studies reporting correlations between increased echo intensity and histologically measured interstitial fat and fibrous tissue in dogs with muscular dystrophy [17], cattle marbling in meat science [16, 17], and individuals with neuromuscular disorders and inflammatory myopathies [18, 19]. These findings have led clinicians and researchers to interpret echo intensity as an accurate index of intramuscular fat and fibrosis in humans and to reflect changes in intramuscular fat and fibrous tissue with ageing, disuse, disease, and exercise.

However, the assumption that echo intensity reflects intramuscular fat and/or fibrous tissue warrants further scrutiny. Fat does not always appear bright (hyperechoic) in sonographic images [20], and studies have reported correlations between echo intensity and intramuscular fat with magnitudes ranging from weak and negative [17, 21] to nearly perfect and positive [22, 23]. Similarly, studies have reported correlations between echo intensity and intramuscular fibrous tissue ranging from weak and negative [18] to nearly perfect and positive [17]. However, these correlations are often based on small, population-specific samples and limited muscle groups, thereby reducing their generalisability. Furthermore, factors unrelated to intramuscular fat or fibrous tissue can also influence echo intensity. For example, transducer-to-muscle distance, which is affected by subcutaneous fat thickness and muscle thickness [23–26], fascicle angle (angle of insonation; [24, 27]), and ultrasound acquisition settings [28] can all alter sound-wave propagation and image brightness and contrast. These factors can affect echo intensity and potentially confound its relationship with intramuscular composition.

For echo intensity to be considered a valid surrogate measure of intramuscular fat and/or fibrous tissue, and therefore a proxy for ‘muscle quality’, several key criteria must be met. The first logical criterion is that echo intensity should demonstrate strong and consistent correlations with intramuscular fat and/or fibrous tissue. Second, these correlations should be generalisable across different muscles and populations. Third, accounting for confounding factors (e.g., pennation angle or transducer-to-muscle distance) should meaningfully influence the observed relationships. If these criteria are met, it would strengthen the case for using echo intensity as a marker of ‘muscle quality’ in both research and clinical settings.

Therefore, the primary aim of this study was to systematically review the literature examining the relationship between ultrasound-derived echo intensity and histological-, chemical-, or MRI-derived measures of intramuscular fat and fibrous tissue, and to quantify the strength of these associations using multi-level meta-analysis to determine whether the available evidence supports ultrasound-derived muscle echo intensity as a valid surrogate marker of ‘muscle quality’. A secondary aim was to assess the consistency of these relationships across muscles and to examine whether methodological and anatomical confounders influence the observed associations.

## 2 METHODS

### 2.1 Literature search strategy

The literature search was conducted in May 2026 in PubMed/MEDLINE, Scopus, Web of Science, EMBASE, EBSCOhost (AMED, APA PsycINFO, CINAHL, and SPORTDiscus), and ProQuest. The searching strategy involved three concept blocks: (i) ultrasound echo-intensity/ echogenicity terms; (ii) intramuscular fat and connective-tissue terms; and (iii) skeletal muscle. Full details of the search strategy, including all keywords and search strings, are presented in Supporting Information Table 1. The search was not limited by publication date. Records were exported and uploaded into Covidence, deduplicated, and screened by two reviewers (MDP and JPN), with conflicts resolved by a third reviewer (RSP). Only full-length original studies were included. Additional studies were identified through backward and forward citation tracking (snowballing) of included studies and targeted searches based on the authors’ knowledge of the literature. These studies underwent the same eligibility and screening procedures as database-derived records. In total, 16 studies were identified through the authors’ knowledge of the literature, of which five were included after screening [29–33]. An updated search was conducted by August 2026, which identified no additional eligible studies. This mixed-approach search strategy is similar to that described by others [34] and has been implemented in previous meta-analyses [35–37]. This review was not prospectively registered.

**Table 1.** Summary of correlations from all meta-analysis models.

| Model | Estimate | Lower CI | Upper CI | Lower PI | Upper PI | No. Effects | No. Studies |
| --- | --- | --- | --- | --- | --- | --- | --- |
| Overall pooled | 0.54 | 0.45 | 0.61 | -0.02 | 0.84 | 204 | 23 |
| Study model |  |  |  |  |  |  |  |
| Animal | 0.46 | 0.22 | 0.64 | -0.43 | 0.89 | 9 | 5 |
| Human | 0.55 | 0.46 | 0.63 | -0.02 | 0.85 | 195 | 19 |
| Population health |  |  |  |  |  |  |  |
| Clinical | 0.64 | 0.42 | 0.79 | -0.01 | 0.91 | 27 | 8 |
| Healthy | 0.49 | 0.39 | 0.56 | -0.09 | 0.82 | 174 | 15 |
| Fat outcome |  |  |  |  |  |  |  |
| IMF | 0.57 | 0.48 | 0.64 | 0.10 | 0.83 | 132 | 22 |
| IMCL | 0.10 | -0.12 | 0.31 | -0.54 | 0.67 | 36 | 4 |
| EMCL | 0.49 | 0.35 | 0.60 | -0.15 | 0.84 | 36 | 4 |
| EI correction |  |  |  |  |  |  |  |
| Raw | 0.53 | 0.44 | 0.61 | -0.04 | 0.84 | 123 | 20 |
| Distance-corrected | 0.57 | 0.46 | 0.66 | -0.05 | 0.87 | 81 | 7 |
| Muscle* |  |  |  |  |  |  |  |
| *Biceps Femoris* | 0.56 | 0.33 | 0.72 | 0.07 | 0.83 | 5 | 2 |
| *Longissimus Dorsi* | 0.41 | 0.16 | 0.61 | -0.12 | 0.76 | 6 | 3 |
| *Supraspinatus* | 0.24 | -0.07 | 0.51 | -0.32 | 0.67 | 12 | 2 |
| *Infraspinatus* | 0.52 | 0.23 | 0.72 | -0.01 | 0.82 | 8 | 2 |
| *Medial Gastrocnemius* | 0.41 | 0.25 | 0.55 | -0.08 | 0.74 | 33 | 4 |
| *Tibialis Anterior* | 0.64 | 0.43 | 0.78 | 0.19 | 0.87 | 4 | 2 |
| *Rectus Femoris* | 0.68 | 0.57 | 0.76 | 0.31 | 0.87 | 21 | 8 |
| *Vastus Lateralis* | 0.53 | 0.38 | 0.65 | 0.07 | 0.80 | 32 | 4 |
| *Vastus Medialis* | 0.43 | 0.26 | 0.57 | -0.06 | 0.75 | 30 | 2 |
| Muscle architecture* |  |  |  |  |  |  |  |
| Pennate | 0.53 | 0.45 | 0.60 | 0.02 | 0.82 | 195 | 22 |
| Non-pennate | 0.63 | 0.44 | 0.77 | 0.13 | 0.88 | 9 | 3 |

IMF=Intramuscular Fat; EMCL=Extramyocellular lipid; IMCL=Intramyocellular lipid
\*Point estimates and precision are not derived from robust models and only shown for categories with $\geq 2$ studies.

### 2.2 Eligibility and data extraction

Manuscripts that met the following criteria were included in the meta-analysis: (a) published in English; (b) published in a peer-reviewed journal; (c) were conducted in skeletal muscles; (d) determined echo intensity from B-mode ultrasonography; (e) reported quantitative echo intensity; (f) measured or estimated intramuscular fat using tissue histology or chemical analysis or magnetic resonance imaging (MRI); (g) measured or estimated intramuscular connective tissue using histology or chemical analysis or MRI; and, (h) reported the correlation between B-mode derived echo intensity and MRI- and/or tissue-derived fat or fibrous tissue. Studies reporting correlations between qualitative MRI fat gradation and echo intensity or reporting qualitative echo intensity classification (Heckmatt scale) were excluded.

The extracted data included sample size, sex, age, health or clinical status, body mass, muscle or muscle group assessed, imaging modality, measurement technique, ultrasound device, and scanning direction. The reference method used to quantify intramuscular fat was classified as chemical analysis, histological analysis, or MRI-based assessment. For MRI-based assessments, techniques were further classified as conventional T1-weighted MRI with signal-intensity-based segmentation, chemical-shift-encoded MRI (CSE-MRI; including Dixon- and IDEAL-based approaches) with water–fat separation, or proton magnetic resonance spectroscopy (^1^H-MRS). The specific fat outcome quantified by each technique (e.g., segmented intramuscular fat/ pixel intensity thresholding, fat fraction, proton-density fat fraction [PDFF], or intra- and extra-myocellular lipid content) was also extracted.

For age and body mass, the overall means and standard deviations were extracted. If a study reported the age for more than one study group, the average age was determined and reported. When standard deviations were not reported, the range was extracted. If a study tested more than one muscle and reported the sample size for the number of subjects of that muscle group, the mean and age of that group was reported.

Studies were classified according to the predominant fascicle architecture of the muscles assessed. Muscles were categorised as either non-pennate, in which fascicles were arranged predominantly parallel to the muscle’s line of action, or pennate, in which fascicles were oriented at an angle to the muscle’s internal tendon or aponeurosis and line of force action [38]. Studies that pooled data across multiple muscles or reported muscle groups were classified as pennate when more than 70% of the assessed muscles had a pennate arrangement. This involved two studies in the echo intensity and intramuscular fat meta-analysis [17, 18], and three studies in the echo intensity and intramuscular fibrosis meta-analysis [refs. 17, 18, 39].

To assess the relationship between echo intensity and intramuscular fat or fibrous tissue, either the Pearson correlation coefficient (*r*) or Spearman’s rank correlation coefficient were extracted. If only the coefficient of determination was reported (R^2^), the square root was computed to determine the Pearson correlation coefficient. When a study reported the correlation coefficient for each muscle group and an overall (pooled) correlation, then the coefficient reported for each muscle was extracted [e.g., 23, 40]. In instances that a study reported only the pooled correlation, then that pooled coefficient was extracted. When studies did not report all correlation values, authors were contacted to request correlation coefficients [31, 41]. For studies in which experimental interventions were employed (e.g., resistance training, fat injection [33], experimentally-induced lesion [42]), only the pre- and/or post-intervention coefficients were extracted, i.e., the correlation between the change scores or within-subject correlations were not extracted. When studies used the same underlying dataset [43–46], the most complete dataset was retained, with duplicate effects excluded and non-overlapping effects retained; publications from the same dataset were assigned the same study identifier[43, 45]. When the studies reported correlation for both the left and right limbs, the data from both limbs were extracted. When studies reported correlations between muscle group fat and muscle echo intensity, the correlation between the primary muscle group and the muscle-specific echo intensity was extracted. For example, Wilkison et al. [32] reported correlations between rectus femoris echo intensity and quadriceps and hamstring fat fraction; in this case, only the correlation between rectus femoris echo intensity and quadriceps fat fraction was extracted. When studies did not report bivariate correlation but reported a predictor-specific P value from a multivariable model, the adjusted partial correlation was derived from that predictor [30, 47]. The corresponding t-statistic was obtained from the reported P value and the model residual degrees of freedom. The partial correlation was then calculated as:

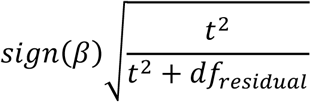

where sign(*β*) indicates the direction of the reported regression coefficient [48]. The residual degrees of freedom were calculated as n – p – 1, where ‘n’ is the sample size and ‘p’ is the number of estimated predictor coefficients, excluding the intercept.

### 2.3 Statistical Analysis

The data spreadsheet is available at the Open Science Framework (10.17605/OSF.IO/GHN96). Effect sizes and the meta-analyses were conducted using R (v 4.5.0 (2025-04-11 ucrt) [49] Vienna, Austria), RStudio (2025.05.0+496, RStudio Team [50], and the ‘metafor’ package (v.5.0.1; [51]).

Correlation coefficients were used as the primary effect size and included both Pearson’s r and Spearman’s rank correlation [52]. Studies reporting predictor-specific adjusted partial correlations from multivariable regression results were also included. Correlation coefficients were transformed to Fisher’s *z* and the sampling variances computed as 1/(*n* − 3), where ‘*n’* represents the sample size, i.e., number of individuals or animals. Pooled estimates, confidence intervals (CI), and prediction intervals (PI) were subsequently back-transformed from Fisher’s *z* to correlation coefficients for interpretation. Prediction intervals represent the expected range of correlations in future studies when no sampling error exists.

Multi-level random-effects meta-analyses were fitted using restricted maximum likelihood estimation with the rma.mv function in the metafor package [51]. Several studies contributed multiple correlation estimates derived from different muscles, outcomes, participant groups, or analytical comparisons. These effect sizes were therefore not treated as fully independent. Random intercepts were specified for studies and individual effect sizes nested within studies (∼1|studyid/effectid), where ‘studyid’ identifies each study and ‘effectid’ identifies each correlation estimate. The study-level random effect represented heterogeneity between studies, whereas the nested effect-level random effect represented residual heterogeneity among effect sizes within studies.

Correlations between the sampling errors of effect sizes within studies were not available; thus, it was not possible to specify the within-study sampling variance–covariance structure. We therefore used cluster-robust variance estimation, clustered at the study level, with small-sample-adjusted robust standard errors, confidence intervals, and statistical tests were performed using ClubSandwich. Heterogeneity was quantified using I² and partitioned into between-study and within-study components using the ‘i2_ml’ function in Orchard 2.0 [53]. Potential small-study effects and publication bias were examined using the funnel plot and the extended Egger-type multilevel meta-regression [54–56].

Several sensitivity analyses were performed. Leave-one-out analysis was performed using the ‘leave_one_out’ function in the ‘orchard 2.0’ package, with models sequentially refitted and the pooled correlation estimated after omitting each study [53, 56]. Additionally, sensitivity analyses addressed two cases in which correlations were calculated from multiple muscles, biopsy samples, or tissue observations obtained from the same individuals [17, 18]. In these studies, the number of observations used to calculate a correlation exceeded the number of independent individuals or animals. Treating all observations as independent could overestimate the effective sample size, underestimate the sampling variance, and consequently assign excessive weight to these studies because observations obtained from the same individual are likely to be highly correlated. Conversely, using only the number of individuals assumes that repeated observations provide no additional independent information and may therefore be conservative. Thus, we ran sensitivity analyses using a range of assumed within-individual intraclass correlation coefficients (ICCs). For each effect size, the effective sample size was estimated as:

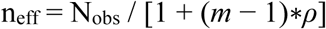

where N_obs_ was the number of muscle or tissue observations used to calculate the correlation, *m* was the mean number of observations per individual, calculated as N_obs_/ N_individuals_, and *ρ* was the assumed correlation among observations obtained from the same individual. The sampling variance was then recalculated as 1/( n_eff_ − 3) when n_eff_ was greater than three. Effect sizes were included in sensitivity models only when n_eff_ was greater than three.

Sensitivity models were assumed within-individual ICCs of 0, 0.2, 0.5, 0.8, and 1.0. An ICC of 0 assumes that all muscle or tissue observations were independent, such that n_eff_ equals the total number of observations, whereas an ICC of 1 assumes that observations obtained from the same individual are perfectly correlated and provide no additional independent information, such that n_eff_ equals the number of individuals. The primary multilevel random-effects structure and study-level cluster-robust inference were applied under each assumption. Pooled and precision estimates were compared across ICC assumptions. Analyses were conducted using all effect sizes for which n_eff_ exceeded three under each assumed ICC.

## 3 RESULTS

A total of 28 eligible studies were included in the study. The earliest study was published in 1993 and the latest in 2026. Two studies reported both fat and fibrosis outcomes [17, 18]. The sample size, sex, age, population health or clinical status, muscle or muscle group assessed, predominant architecture, imaging modality, measurement technique, ultrasound device and scanning direction, whether ultrasound imaging correction was applied, reference method used to quantify intramuscular fat, and the specific technique used within each reference method are reported in Supporting Information, Table 2. The systematic search and study-selection process flowchart is shown in Figure 1.

**Figure 1.**
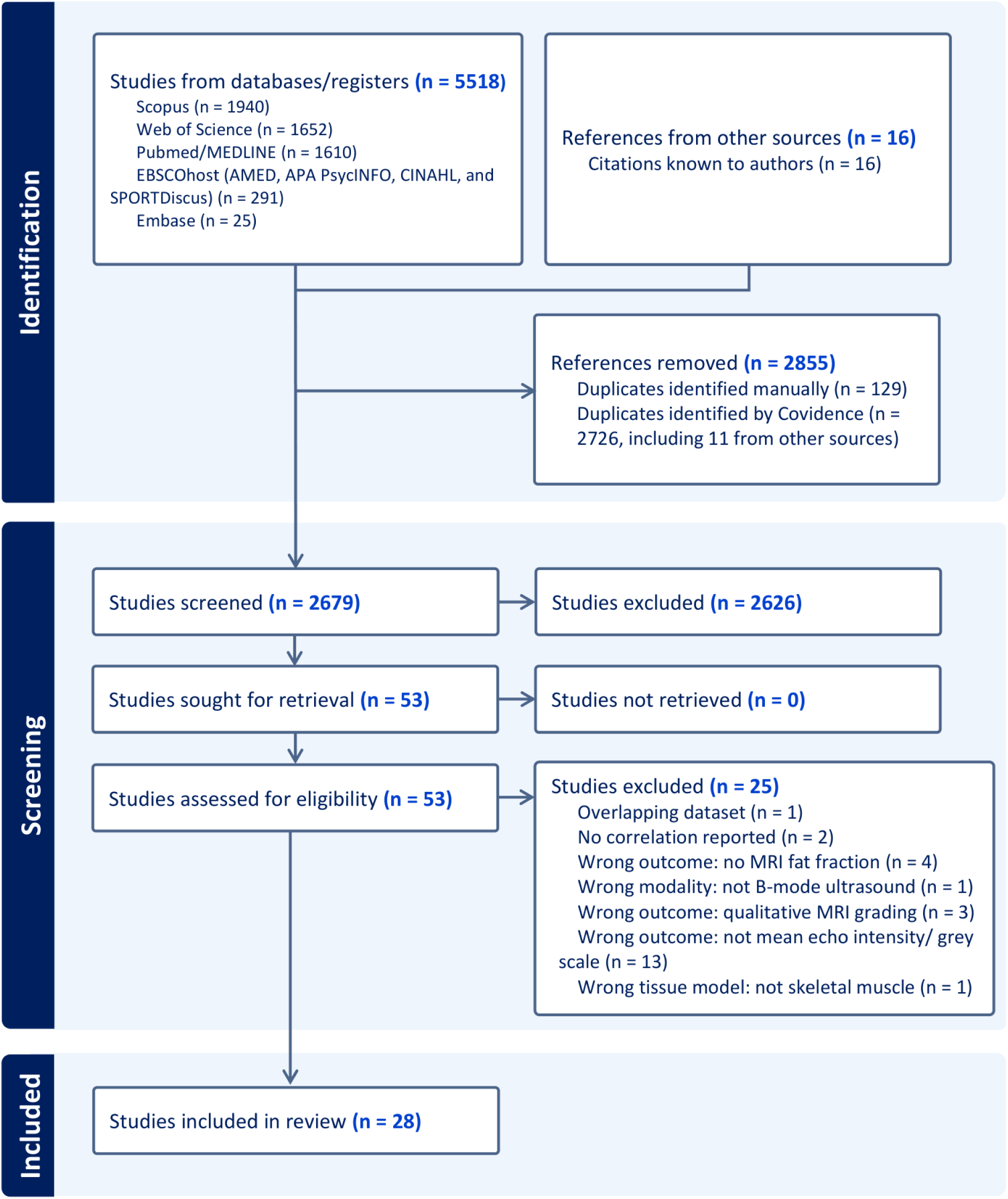
PRISMA (Preferred Reporting Items for Systematic Reviews and Meta-Analyses) flow chart.

Of the 28 eligible studies, 24 contributed 205 effect sizes to the meta-analysis examining the association between ultrasound mean echo intensity and intramuscular fat [17, 18, 23, 30–33, 41, 43, 45, 57–67]. One study was excluded from the primary analysis because correlations were calculated from multiple muscle or tissue observations obtained from only two animals [17], resulting in 23 independent studies and 204 effect sizes. When the number of independent animals was used as the sample size, the sampling variance of Fisher’s z could not be estimated because the sample size was n≤3. Seven studies reported associations between distance-corrected mean echo intensity and intramuscular fat, contributing 81 effect sizes [23, 29, 30, 41, 57, 66, 68]. Four of these studies also reported uncorrected echo intensity, contributing 60 effect sizes for each measure [23, 57, 66, 68].

Five studies were eligible for the meta-analysis examining the association between ultrasound echo intensity and intramuscular fibrous tissue [17, 18, 39, 42, 69]. These studies contributed to 7 effect sizes, although one study was excluded because the number of independent animals used as the sample size was ≤ 3, totalling 6 effects for the main analysis. One study contributed two effects for associations between distance-corrected echo intensity and intramuscular fibrosis [69].

### 3.1 Echo intensity─intramuscular fat relationship

#### 3.1.1 Main model

The multilevel meta-analysis included 204 effect sizes from 23 studies. The pooled correlation was 0.54 (95% CI [0.45, 0.61], 95% PI [−0.02, 0.84]; P<0.01; Figure 2). Total heterogeneity was substantial (I²=74.5%), with similar contributions from between-study (I²_study_=40.4%) and within-study variations (I²_study/effectid_=34.1%). The wide prediction interval suggests that the true correlation in a comparable population could range from negligible to strong. Visual inspection of the funnel plot did not suggest asymmetry (Figure 1, Supporting Information), and the Egger-type meta-regression did not provide clear evidence of an association between effect size and its standard error, suggesting no small-study effects (F_(1, 7.10)_=1.64, P=0.24). Leave-one-out and sensitivity analyses across the prespecified range of assumed within-individual intraclass correlation coefficients did not meaningfully alter the results (see Figure 2 and Table 3 in Supporting Information).

**Figure 2.**
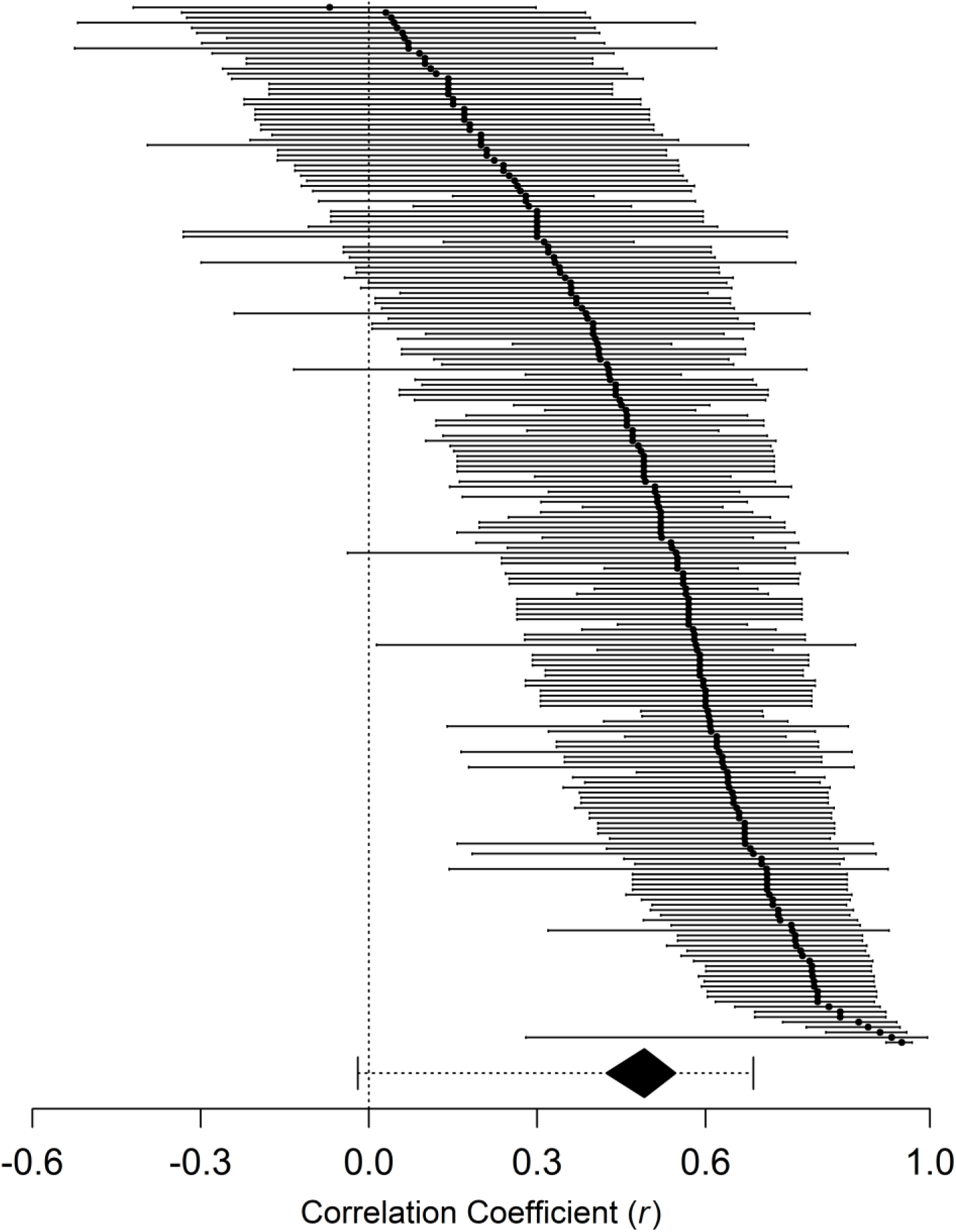
Caterpillar plot of individual correlation coefficients of the relationship between ultrasound echo intensity and intramuscular fat measures using MRI or histological or histochemical analysis across all 204 effects in the 23 studies included in the meta-analysis. Each circle represents an individual effect estimate, with horizontal lines indicating the 95% confidence interval (CI). The dashed vertical line at 0 represents no correlation. Values to the right of 0 indicate a positive association between echo intensity and intramuscular fat, whereas values to the left indicate a negative association. The pooled robust random-effects estimate is shown by the black diamond, with its width representing the 95% CI; the dashed horizontal interval around the pooled estimate represents the 95% prediction interval (PI). Estimates were analysed on the Fisher’s z scale and back-transformed to display as correlation coefficients. The pooled correlation was 0.54 (95% CI [0.45, 0.61], 95% PI [−0.02, 0.84].

#### 3.1.2 Moderators

Table 1 summarises the correlations from all meta-analytical models, including the overall and exploratory analysis of moderator effects.

##### Study model

We explored whether the population studied was a moderator of the association between echo intensity and intramuscular fat. The pooled correlation was 0.46 for animal studies (95% CI [0.22, 0.64]; 95% PI [-0.43, 0.89]) and 0.55 for human studies (95% CI [0.46, 0.63], 95% PI [-0.02, 0.85]). However, cluster-robust inference provided no clear evidence that the association differed between human and animal studies (F_(1,4.40)_=1.68, P=0.26). Residual heterogeneity remained substantial (I^2^=74.8%), with similar contributions from between-study (I^2^_study_=41.4%) and within-study variations (I^2^_study/effect_=33.4%). The wide prediction interval for animal studies indicated considerable uncertainty in the correlation expected across animal populations and muscles. These findings should be interpreted cautiously because relatively few animal studies were available and the numbers of studies and effect sizes were highly unbalanced between study models (Table 1), reducing the precision of the cluster-robust inference.

##### Population health status

We explored whether population health status moderated the association between echo intensity and intramuscular fat. The pooled correlation was 0.64 for clinical populations (95% CI [0.42, 0.79]; 95% PI [-0.01, 0.91]) and 0.49 for healthy populations (95% CI [0.39, 0.56]; 95% PI [-0.09, 0.82]). Cluster-robust inference provided no clear evidence that the association differed among population health categories (F_(1, 9.53)_=3.04, P=0.11). Residual heterogeneity remained substantial (I^2^=72.8%), with similar contributions from between-study (I^2^_study_=37.3%) and within-study variations (I^2^_study/effect_=35.5%). The wide confidence and prediction intervals indicated considerable uncertainty in the expected association and should therefore be interpreted cautiously because the numbers of studies and effect sizes were unbalanced between the population health categories, resulting in limited precision for the moderator test and estimates (Table 1). Sensitivity analysis including the mixed cohorts similarly provided no clear evidence of moderation by population health status (F_(2, 1.96)_=1.00, P=0.50).

##### Intramuscular fat outcome

We also examined muscle fat compartment (intra-or extra-myocellular lipid, or total intramuscular fat) as a moderator of the association between echo intensity and intramuscular fat. The pooled correlation was 0.57 for total intramuscular fat (95% CI [0.48, 0.64]; 95% PI [0.10, 0.83]), 0.10 for intramyocellular lipid (95% CI [-0.12, 0.31]; 95% PI [-0.54, 0.67]), and 0.49 for extramyocellular lipid (95% CI [0.36, 0.60]; 95% PI [-0.15, 0.83]). Analyses revealed a significant moderation effect of fat outcome (F_(2, 201)_=65.1, P<0.001). However, cluster-robust variance estimation revealed no significant moderation effect (F_(2, 0.22)_ = 69.1, P = 0.50). Given the small number of independent studies, the cluster-robust estimate was considered less informative, and we thus used model-based estimates for interpretation. Residual heterogeneity remained substantial (I²=69.1%) and was primarily attributable to between-study variation (I²_study_=58.2%), with a smaller contribution from within-study variation (I²_study/effect_=10.8%).

##### Effect of distance correction

Correction of echo intensity for the distance between the skin and muscle was examined as a moderator of the association between echo intensity and intramuscular fat. The pooled correlation was 0.53 for raw echo intensity (95% CI [0.44, 0.61]; 95% PI [−0.04, 0.87]) and 0.57 for distance-corrected echo intensity (95% CI [0.46, 0.66]; 95% PI [−0.05, 0.87]). Cluster-robust inference provided no clear evidence that the association differed between raw and corrected estimates (F_(1, 1.74)_=2.92, P=0.25). Residual heterogeneity remained substantial (I²=74.9%), with similar contributions from between-study (I²_study_=42.1%) and within-study variations (I²_study/effect_=32.8%).

We performed an exploratory head-to-head analysis from studies reporting correlations between intramuscular fat and both raw and distance-corrected echo intensity. The pooled correlation was 0.57 for raw echo intensity (95% CI [0.29, 0.76]; 95% PI [−0.40, 0.94]) and 0.61 for distance-corrected echo intensity (95% CI [−0.30, 0.80]; 95% PI [−0.36, 0.95]). Cluster-robust inference provided no clear evidence that the correlations differed between raw and distance-corrected echo intensity (F_(1, 1.42)_=5.11, P=0.20). Residual heterogeneity remained substantial (I²=72.3%), with similar contributions from within-study (I²_study/effect_=40.7%) and between-study variation (I²_study_=31.6%). These findings should be interpreted with caution because only four independent studies contributed to the analysis and one study contributed 80% of the effects.

##### Muscle

We explored whether the muscle assessed moderated the association between echo intensity and intramuscular fat. Model-based inference revealed that the association differed across muscles (F_(21, 182)_=3.29, P<0.001). However, the model included 22 muscle categories across only 23 independent studies, and several categories were represented by a single study. Consequently, a test of moderation effect using cluster-robust inference could not be obtained, and cluster-robust estimates for some muscle categories were unstable and not reported. Residual heterogeneity remained substantial (I²=67.2%), with most variance from between-study variation (I²_study_=42.0%) than within-study variation (I²_study/effect_=25.2%).

##### Muscle architecture

We also explored whether muscle architecture moderated the association between echo intensity and intramuscular fat. The pooled correlation was 0.63 for non-pennate muscles (95% CI [0.44, 0.77]; 95% PI [0.13, 0.88]) and 0.51 for pennate muscles (95% CI [0.45, 0.60]; 95% PI [0.02, 0.82]). There was no clear moderation effect of architecture in the correlation between echo intensity and intramuscular fat [(F_(1, 202)_=1.32, P=0.25); and cluster-robust inference (F_(1, 2.14)_=0.47, P=0.56)], likely because the non-pennate group had only nine effects from three studies. The cluster-robust 95% CI and 95% PI for the non-pennate group were −0.26 to 0.94 and −0.68 to 0.98, whereas those for pennate muscles were 0.44 to 0.60 and -0.02 to 0.83, respectively. Residual heterogeneity remained substantial (I²=73.1%), with similar contribution from between-study (I²_study_=36.4%) and within-study variations (I²_study/effect_=36.7%).

### 3.3 Echo intensity─intramuscular fibrous tissue relationship

#### 3.3.1. Main Model

The multilevel meta-analysis included six effect sizes from four studies. The pooled correlation between echo intensity and intramuscular fibrous tissue was 0.34 (95% CI [−0.39, 0.81]; 95% PI [−0.85, 0.96], P=0.24; Figure 3). Total heterogeneity was substantial (I²=87.9%) and predominantly from between-study variation (I²_study_=87.9%). The wide confidence and prediction intervals indicate substantial uncertainty, with the correlation expected in a comparable population potentially ranging from strongly negative to strongly positive. These findings should be interpreted cautiously because the analysis included only four independent studies. Sensitivity analyses across the prespecified range of assumed within-individual intraclass correlation coefficients did not alter the findings; the pooled correlations ranged from 0.34 to 0.50 and were not statistically significant (Table 4, Supporting Information).

**Figure 3.**
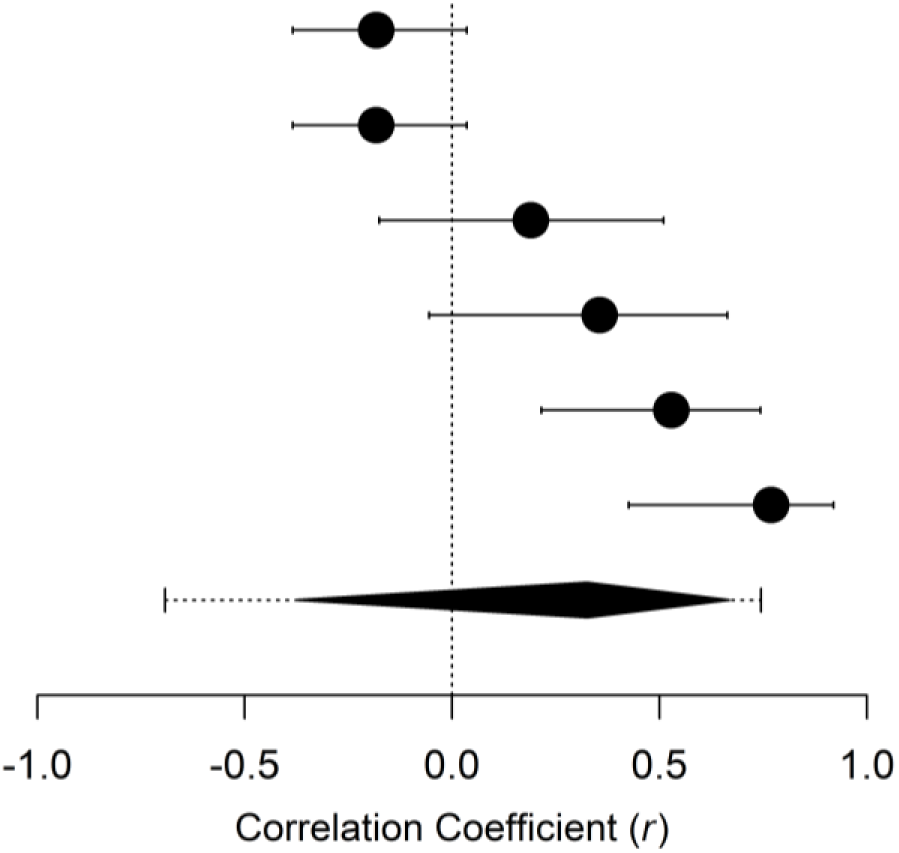
Caterpillar plot of individual correlation coefficients for the relationship between ultrasound echo intensity and intramuscular fibrous tissue across six effect estimates from four studies included in the meta-analysis. Each circle represents an individual effect estimate, with horizontal lines indicating the 95% confidence interval (CI). The dashed vertical line at 0 represents no correlation; values to the right indicate a positive association between echo intensity and intramuscular fibrous tissue, whereas values to the left indicate a negative association. The cluster-robust pooled multilevel estimate is shown by the black diamond, with its width representing the 95% CI; the dashed horizontal interval represents the 95% prediction interval (PI). Estimates were analysed on the Fisher’s *z* scale and back-transformed to display as correlation coefficients. The pooled correlation was 0.34 (95% CI [−0.39, 0.81]; 95% PI [−0.85, 0.96]).

## 4. DISCUSSION

The primary aim of this systematic review and meta-analysis was to examine the relationship between ultrasound-derived echo intensity and reference measures of intramuscular fat and fibrous tissue. We find that echo intensity is weakly-to-moderately associated with intramuscular fat, and the magnitude of this association varied considerably across studies and muscles. In contrast, we find that echo intensity does not appear to be associated with intramuscular fibrous tissue. Exploratory analysis indicates that the relationship between echo intensity and intramuscular fat is moderated by muscle assessed and may be influenced by fat compartment (intracellular vs. extracellular). However, this relationship does not seem to be influenced by study model (animal vs. human), population health status (healthy vs. clinical), probe-to-muscle distance correction (corrected vs. not corrected), or muscle architecture (non-pennate vs. pennate).

### 4.1 Relationship between echo intensity and intramuscular fat

In the present study, we synthesise 204 effects from 23 independent studies to provide the most comprehensive evaluation to date of the relationship between ultrasound-derived echo intensity and intramuscular fat. We find that echo intensity was significantly associated with intramuscular fat, but that the magnitude of this correlation was weak-to-moderate (*r*=0.54). This finding provides some support to the long-standing premise that greater intramuscular fat infiltration contributes to increased echo intensity [15], but also suggests that intramuscular fat accounts for only part of the variation in echo intensity. This premise was initially based on studies reporting correlations between histology-derived estimates of intramuscular fat or lipid content and ultrasound-derived echo intensity in dogs with muscular dystrophy [17], cattle marbling [16, 17], and individuals with neuromuscular disorders [18, 19], and was subsequently investigated in humans using non-invasive estimates from MRI [e.g., 23, 62, 65]. However, these studies have generally included small samples, and the reported correlations have varied substantially, ranging from weak and negative [17, 21] to nearly perfect and positive [22, 23]. The current findings confirm this considerable between-study variability. The 95% prediction interval in the present study ranged from −0.02 to 0.84, indicating that the true association between echo intensity and intramuscular fat in a future study could plausibly range from negligible to very strong. These findings provide insufficient evidence to support the use of echo intensity as an accurate surrogate measure of intramuscular fat.

#### 4.1.1 Intramuscular fat reference method and compartment

We expected the association between echo intensity and intramuscular fat to vary with fat reference method because different methods characterise different aspects of intramuscular fat. Histological methods allow quantification of interstitial fat within muscle sections, whereas chemical extraction or biochemical assays allow quantification of total extractable fat or triglyceride content. MRI-derived intramuscular fat may also vary with acquisition and quantification approaches. Studies used T1-weighted imaging and chemical-shift Dixon techniques with differing acquisition parameters and fat-water signal modelling that can affect fat estimates [70] and may have contributed to the substantial heterogeneity observed. Fat compartment was also a plausible moderator because MR spectroscopy can distinguish intramyocellular from extramyocellular lipid [71–73] and previous evidence suggests that echo intensity is more strongly associated with lipid stored outside than inside muscle fibres [62]. Consistent with this, subgroup analyses revealed a weak-to-moderate association with extramyocellular lipid but only a weak association with intramyocellular lipid (see Table 1). This may reflect the larger or more numerous acoustic interfaces created by extramyocellular lipid than microscopic lipid droplets within muscle fibres. However, the association with extramyocellular lipid was not stronger than that with total intramuscular fat, and cluster-robust inference for the moderator effect was inconclusive because only four studies examined intra-and extra-myocellular lipid concentrations [31, 62, 64, 66]. Thus, the findings suggest that the fat compartment may influence the association between echo intensity and intramuscular fat, particularly for extramyocellular lipid, but this should be interpreted cautiously given the small number of studies.

#### 4.1.2 Muscle-specific variation and muscle architecture

Numerous studies have shown that both intramuscular fat and echo intensity vary substantially among muscles [25, 74, 75] and previous work suggests that their association is also muscle-dependent [23]. Consistent with this, the strength of the association significantly varied across muscles in the current analyses (Table 1), indicating that relationships observed in one muscle may not generalise to others. Muscle architecture may partly explain this variation because echo intensity is substantially affected by fascicle orientation due to direction-dependent (anisotropic) effects [24, 27]. Nonetheless, the correlation between intramuscular fat and echo intensity was not substantially stronger in non-pennate than pennate muscles (0.63 vs. 0.53), and this difference was not clearly supported by moderation analyses, likely because estimates from non-pennate muscle involved only nine effects from three independent studies. Nevertheless, the influence of fascicle angle is mechanistically plausible because the energy of sound waves reflected back to the transducer depends on the orientation of the fascicles relative to the ultrasound beam [24, 27]. We previously showed that experimentally reducing *vastus lateralis* fascicle angle increased echo intensity and that changing insonation angle through beam steering markedly changed echo intensity [24, 27]. Thus, differences in fascicle angle among muscles and participants, as well as small variations in probe or limb position, could alter echo intensity independently of tissue composition and contribute to variation in its association with intramuscular fat. These muscle-specific differences should therefore not be interpreted as evidence that echo intensity is inherently valid in some muscles; rather, they identify muscle architecture as a plausible source of measurement variability that warrants further investigation.

#### 4.1.3 Influence of methodological factors and study characteristics

Subcutaneous fat thickness and muscle depth have been proposed as important confounders of echo intensity [23, 26] because attenuation by overlying tissues and greater propagation distance can reduce the returned ultrasound signal independently of intramuscular fat. Mathematical correction procedures have therefore been proposed to account for muscle-to-transducer distance with the expectation that it would strengthen the relationship between echo intensity and intramuscular fat. However, correlations did not differ between studies using distance-corrected (r=0.57) and uncorrected (r=0.53) echo intensity, with consistent findings in head-to-head analyses. This does not suggest that subcutaneous fat thickness and/or composition or muscle depth are unimportant; rather we hypothesise that correction factors may systematically alter echo intensity without substantially altering participant ranking and, consequently, the correlation coefficient. Existing equations also primarily account for subcutaneous fat thickness or depth and may not capture variation in skin and subcutaneous fat composition that may additionally affect attenuation and reflection independently of distance [24, 76]. Thus, while the physical rationale is strong, current correction procedures do not appear to significantly strengthen the association between echo intensity and intramuscular fat.

### 4.2 Relationship between echo intensity and intramuscular fibrous tissue

In contrast to the observed relationship between echo intensity and intramuscular fat, we found no clear evidence that echo intensity is associated with intramuscular fibrous tissue. The pooled correlation was weak (r=0.34), with substantial uncertainty because only four studies contributed to six effects. This was somewhat unexpected because echo intensity is commonly proposed to reflect intramuscular fibrosis, and a strong association with histologically quantified fibrous tissue has been reported in a canine model of muscular dystrophy [17]. However, the included studies predominantly quantified the amount or area fraction of collagen, fibrous tissue, or broader intramuscular connective tissue as histological measures of fibrosis [17, 18, 39, 42, 69], whereas ultrasound backscatter and the intensity of echoes returned may also depend on the organisation and orientation of collagenous structures [77]. Indeed, fibrosis involves collagen deposition as well as changes in collagen architecture and cross-linking [78]. Supporting this, evidence suggests that experimentally altering collagen organisation and cross-linking in rat myocardium changed ultrasonic backscatter (brightness) despite no detectable change in collagen content [79], while changes in the three-dimensional structural organisation in muscle-mimicking phantoms similarly altered echo intensity [80]. Thus, variation in collagen organisation and orientation may partly explain the inconsistent relationship between echo intensity and fibrous tissue. Overall, current evidence is insufficient to support echo intensity as a valid surrogate of intramuscular fibrosis, although the limited evidence available precludes excluding an influence of fibrosis on echo intensity.

### 4.3 Considerations for interpreting echo intensity as an index of ‘muscle quality’

Ageing, disuse, and neuromuscular disorders are often associated with increased intramuscular fat infiltration and fibrous tissue accumulation [3, 4], reducing ‘muscle quality’. Ultrasound-derived echo intensity has been extensively used as a surrogate measure of intramuscular fat and fibrosis and, consequently, to track these changes in muscle quality. However, for echo intensity to be a valid surrogate or index of intramuscular fat or fibrosis, it should demonstrate strong associations with validated reference measures, these associations should be generalisable across muscles and populations, and they should strengthen when known confounding factors are accounted for. The current results indicate that the association between echo intensity and intramuscular fat is weak-to-moderate, varies largely across muscles, and does not seem to be significantly strengthened by current correction factors, while evidence for its association with intramuscular fibrosis is weak. Therefore, current evidence does not immediately support echo intensity as an accurate surrogate measure or an index of intramuscular fat or fibrosis and, therefore, of ‘muscle quality’. This is important because several studies label echo intensity scores as intramuscular adipose tissue [81–86], and commercially available ultrasound systems such as ‘MuscleSound’ advocate estimating intramuscular fat percentage using equations adapted from a single study and developed for a single muscle [23, 87]. The heterogeneity observed in the present study suggests that such associations observed in one muscle should not be generalised to others.

The evidence presented herein is derived from cross-sectional studies and does not allow establishing whether changes in echo intensity track changes in intramuscular fat and/or fibrosis with ageing or in response to (non)pharmacological therapies. Experimental evidence shows that serial fat injection into *ex vivo* bovine tongue exhibited a dose-dependent increase in echo intensity (within-subject correlation of 0.93, 95% CI [0.80, 0.98]). However, this involved manipulating a single variable in a muscle with relatively simple architecture. In humans, changes in hydration, oedema, inflammation, muscle size, fascicle angle, and overlying tissues may confound intervention-related changes in echo intensity. Future studies should therefore account for these potential confounders and examine longitudinal changes in echo intensity against reference measures or determine whether other quantitative ultrasound approaches targeting intrinsic acoustic tissue properties [88] or texture-based analyses [16, 89, 90] may alternatively provide more specific measures of muscle composition and their changes.

### 4.4 Limitations

The current study has limitations. First, ultrasound systems, acquisition settings, image processing, and reference methods varied across studies and potentially contributed to between-study heterogeneity. Second, association rather than agreement was assessed, precluding evaluation of interchangeability between echo intensity and reference measures. Thus, even a strong association might indicate that higher echo intensity accompanies greater intramuscular fat or fibrous tissue, but not that it accurately quantifies either tissue component. Third, multiple effects from the same studies or individuals required assumptions about within-study dependence, although sensitivity analyses did not significantly alter the pooled estimates. Fourth, moderator and subgroup analyses were limited by the small number of independent studies and should be interpreted cautiously. Finally, no formal risk-of-bias or certainty-of-evidence assessment was performed.

## 5 Conclusion

The results of the present study demonstrate that echo intensity is weakly-to-moderately associated with intramuscular fat, with substantial variation across studies and muscles, and that current correction procedures do not seem to meaningfully strengthen this relationship. The association between echo intensity and intramuscular fibrous tissue was limited and uncertain. Thus, the current evidence provides limited support for interpreting echo intensity as a surrogate measure of intramuscular fat or fibrosis and, on this basis, as an index of ‘muscle quality’.

## Data Availability

The dataset is available online in OSF

https://doi.org/10.17605/OSF.IO/GHN96

## Ethical approval

Not applicable.

## Data availability

Data extraction dataset is available through the Open Science Framework - 10.17605/OSF.IO/GHN96.

## Ethical Guidelines Statement

All authors of this manuscript comply with the guidelines of ethical authorship and publishing in the Journal of Cachexia, Sarcopenia and Muscle.

## SUPPORTING INFORMATION

**Supporting Information Figure 1.**
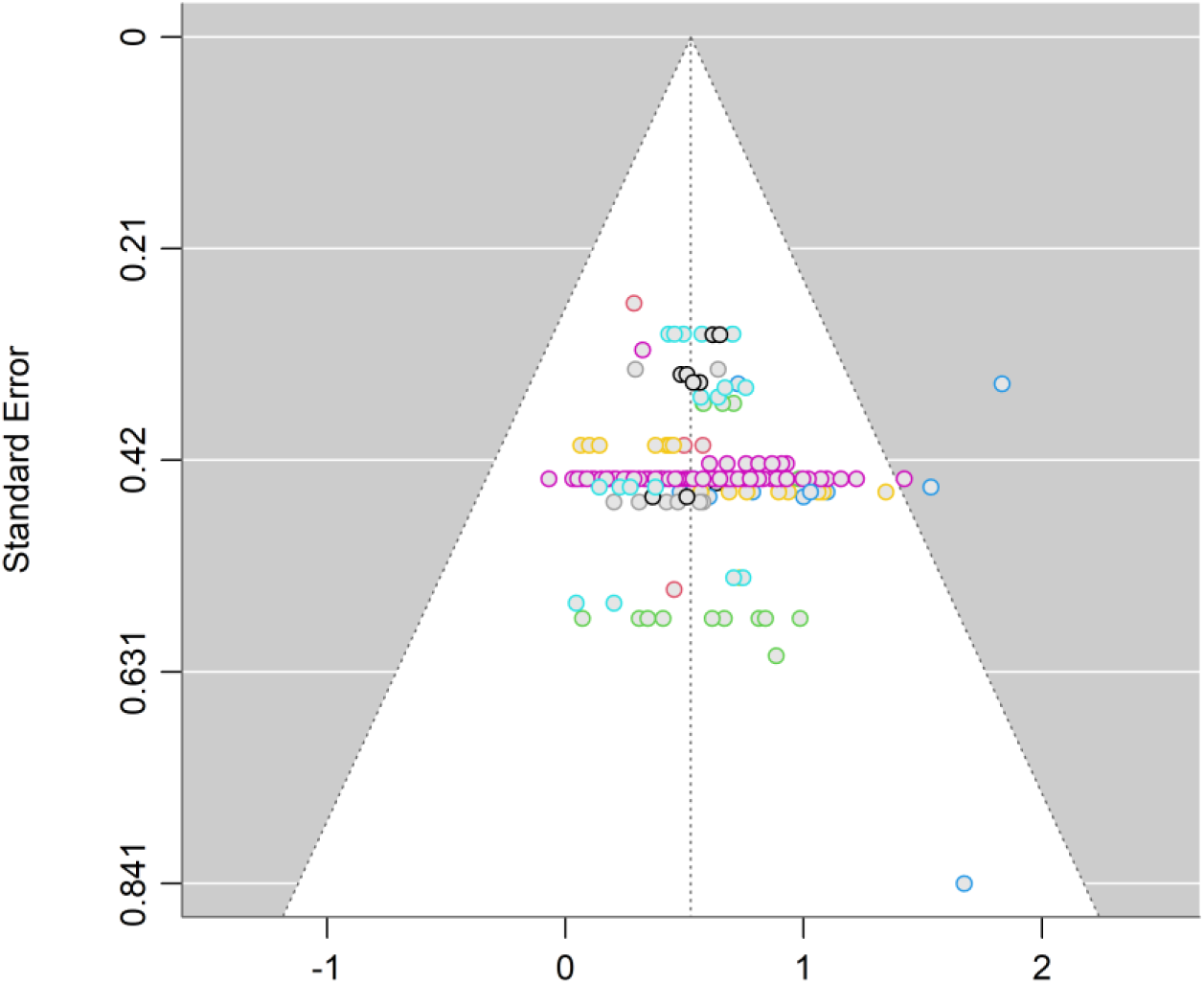
Funnel plot of the Fisher’s z-transformed correlation. Individual studies are coloured.

**Supporting Information Figure 2.**
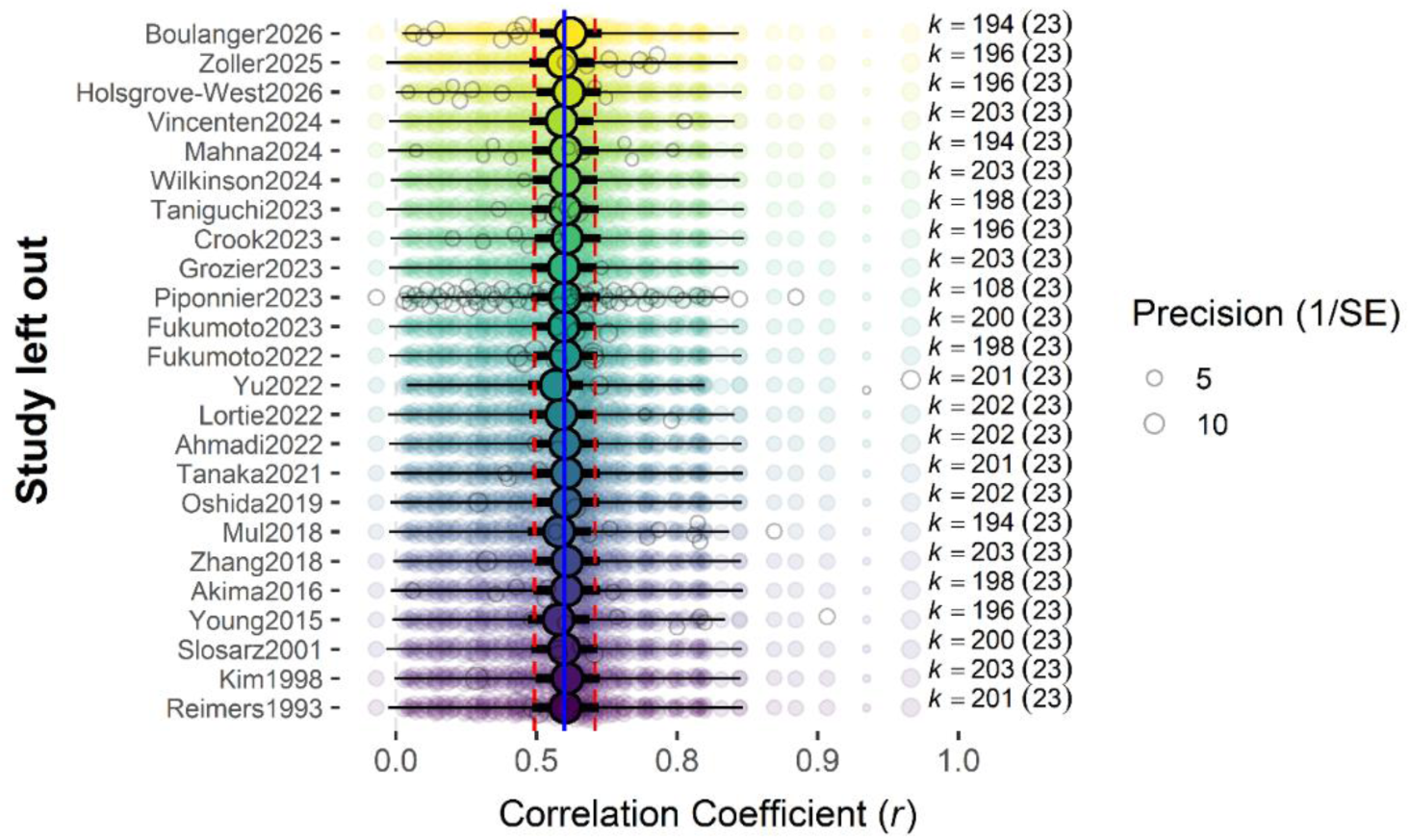
Leave-one-out analysis. For each row, the multilevel meta-analytic model is refitted after removing the indicated study (“Study left out” on the y-axis). Semi-transparent coloured points show the individual effect sizes contributing to each refitted model, with point size proportional to precision (1/standard error (SE); legend). Empty grey circles indicate the effect sizes omitted from that refit. The coloured circles indicate the pooled coefficient of correlation estimate from each leave-one-out refit. The thick black horizontal lines show the corresponding 95% confidence interval (CI), and the thin black horizontal lines show the corresponding 95% prediction interval (PI). The solid blue vertical line indicates the pooled estimate from the full dataset (r=0.54), and the red dashed vertical lines indicate the 95% CI of the overall effect in the original full model. k represents the number of effect sizes remaining in each refit, with the number of studies shown in parentheses. Removing the two most influential studies yielded back-transformed correlations of 0.51 (95% CI [0.44, 0.58], 95% PI [0.03, 0.80]) and 0.55 (95% CI [0.47, 0.62], 95% PI [0.02, 0.84]).

**Table 1.**
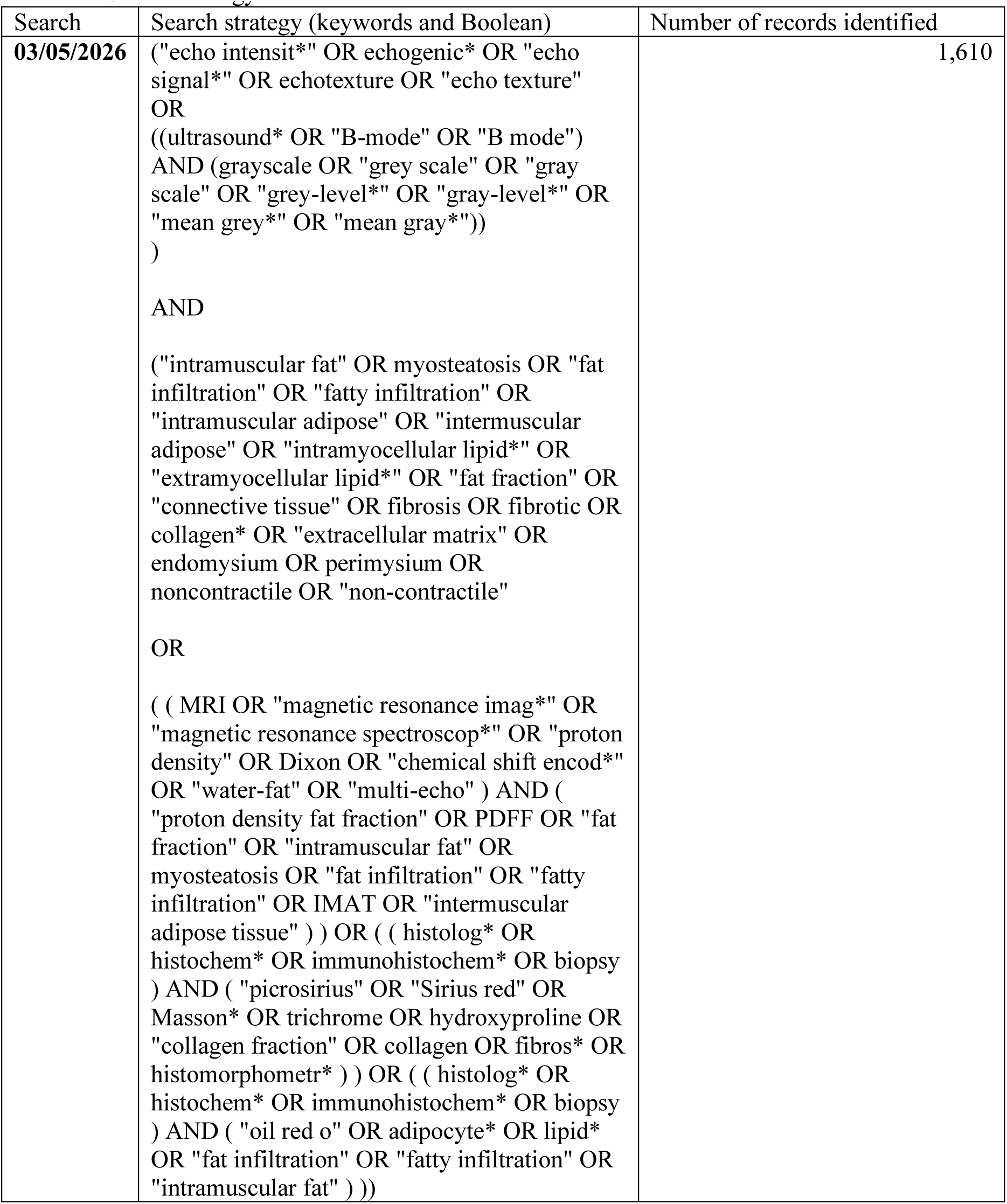
Search strategy results.

**Table 2.**
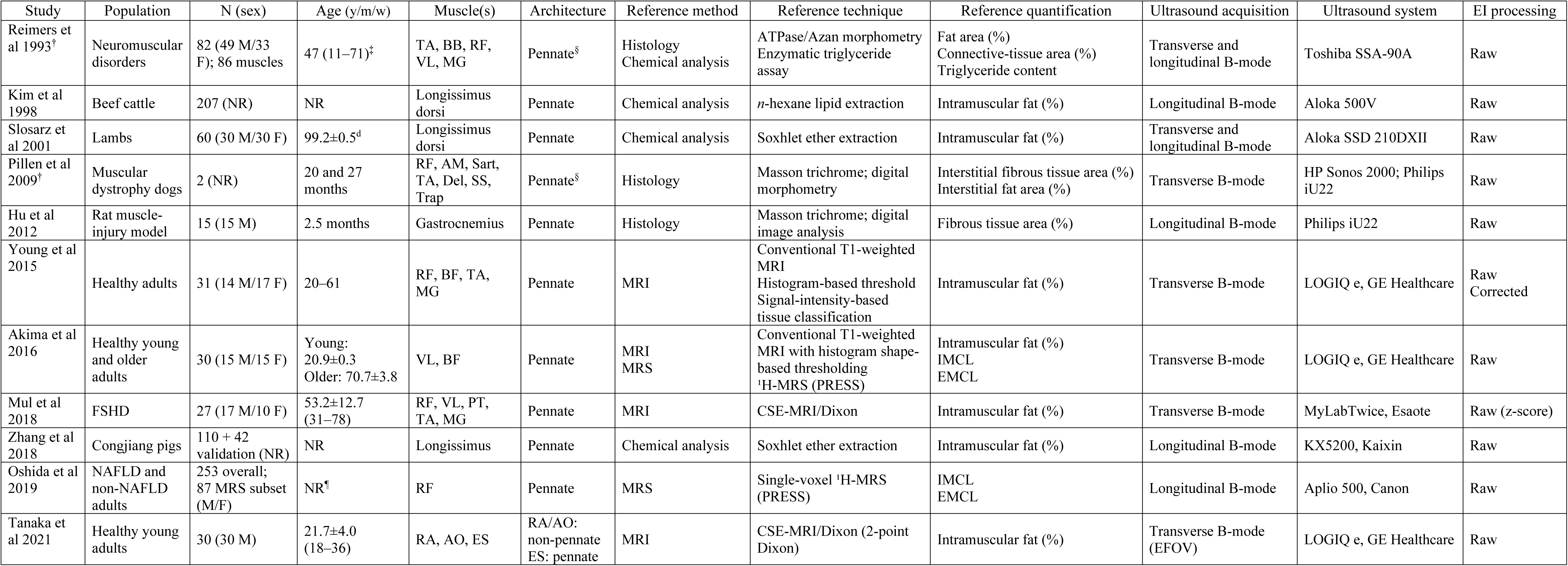
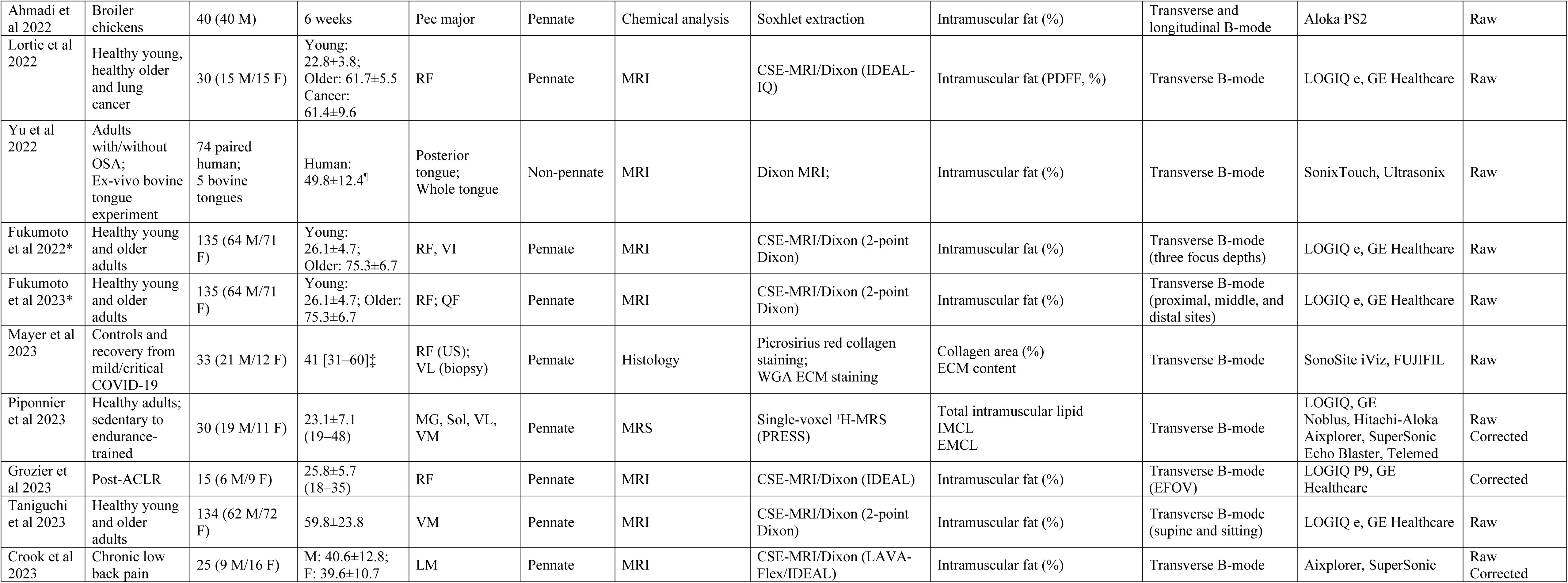
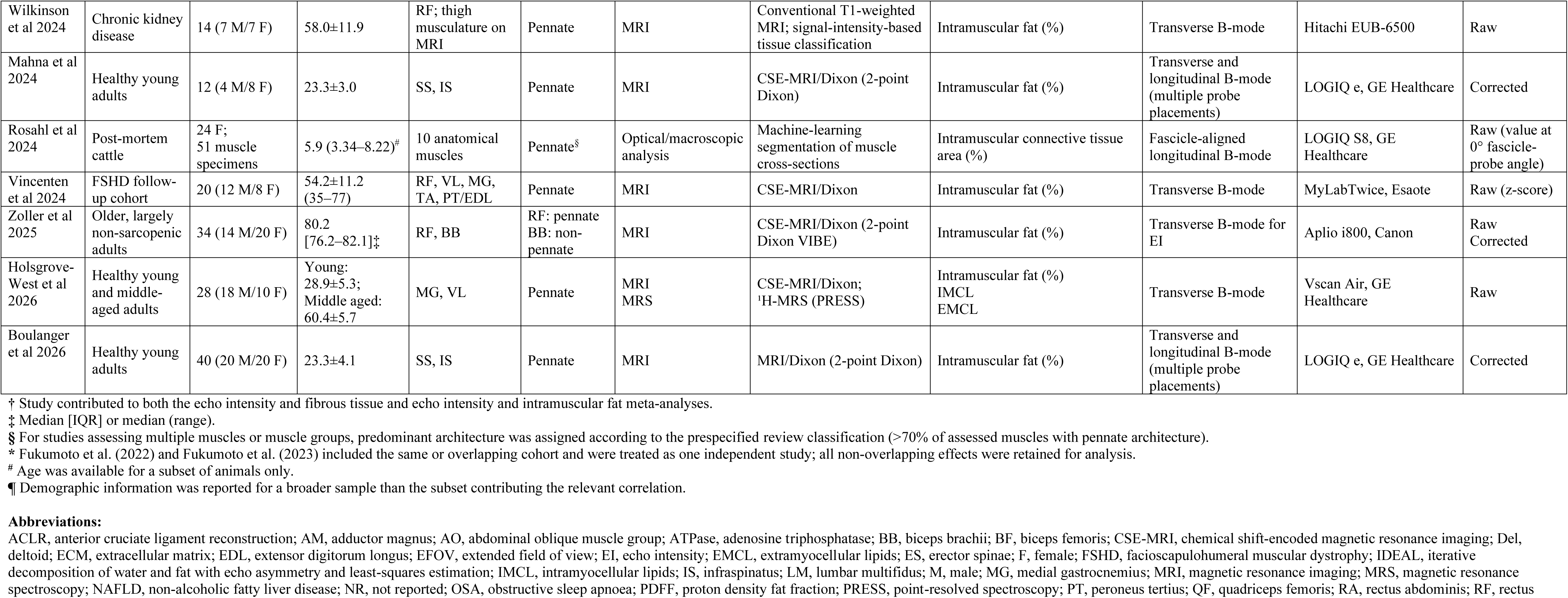
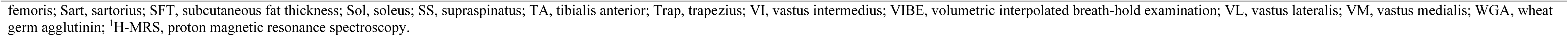
Characteristics of studies evaluating B-mode ultrasound echo intensity and reference measures of intramuscular fat and fibrosis/connective tissue.

**Supporting Information Table 3.**
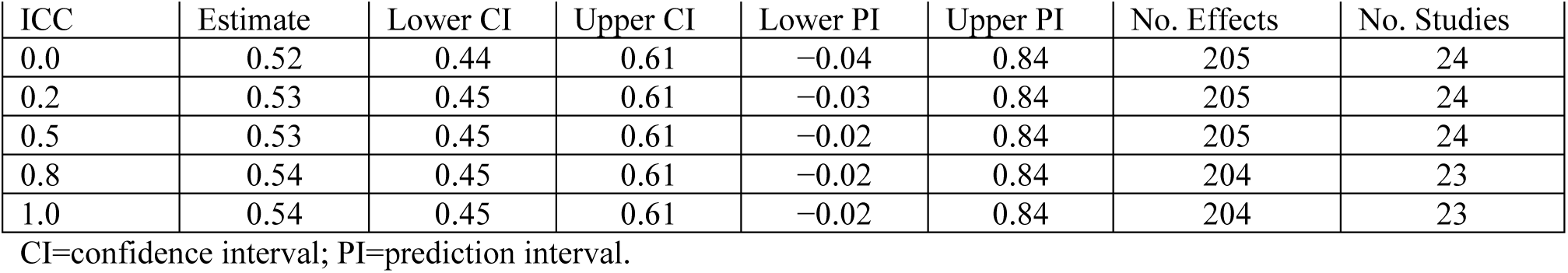
Sensitivity analysis of the pooled correlation between ultrasound-derived echo intensity and measures of intramuscular fat tissue using pre-specified assumptions about the dependence of observations within individuals, i.e., within-individual correlations (ICC). Results are shown as robust estimates with Pearson’s correlation coefficients back-transformed from Fisher’s z.

**Supporting Information Table 4.** Sensitivity analysis of the pooled correlation between ultrasound-derived echo intensity and measures of intramuscular fibrous tissue using pre-specified assumptions about the dependence of observations within individuals, i.e., within-individual correlations (ICC). Results are shown as robust estimates with Pearson’s correlation coefficients back-transformed from Fisher’s z.

| ICC | Estimate | Lower CI | Upper CI | Lower PI | Upper PI | No. Effects | No. Studies |
| --- | --- | --- | --- | --- | --- | --- | --- |
| 0.0 | 0.50 | -0.19 | 0.86 | -0.83 | 0.98 | 7 | 5 |
| 0.2 | 0.45 | -0.21 | 0.83 | -0.80 | 0.97 | 7 | 5 |
| 0.5 | 0.36 | -0.32 | 0.80 | -0.81 | 0.96 | 7 | 5 |
| 0.8 | 0.34 | -0.39 | 0.81 | -0.85 | 0.96 | 6 | 4 |
| 1.0 | 0.34 | -0.39 | 0.81 | -0.85 | 0.96 | 6 | 4 |

CI=confidence interval; PI=prediction interval.

